# White Matter Integrity Differences in 2-year-old Children Treated with ECMO: A Diffusion-Weighted Imaging Study

**DOI:** 10.1101/2024.03.11.24304090

**Authors:** Michaela Ruttorf, Julia Filip, Thomas Schaible, Meike Weis, Frank G. Zöllner

## Abstract

School-aged and adolescent survivors of neonatal extracorporeal membrane oxygenation (ECMO) treatment still suffer from neurodevelopmental delays such as verbal, visuo-spatial and working memory problems, motor dysfunction and sensorineural hearing loss, respectively, later in life. These neurodevelopmental delays are normally assessed by neuropsychological testing within follow-up programs. The purpose of this study is to demonstrate that diffusion-weighted imaging (DWI) in 2-year-old survivors of neonatal ECMO treatment might be a predictor of neurodevelopmental outcome. Therefore, 56 children underwent DWI at 3 T. Fractional anisotropy (FA), first fibre partial volume fraction estimate (F1) and radial diffusivity (RD) are compared using tract-based spatial statistics adapted to a paediatric brain atlas and whole-brain voxelwise statistics with age and gender as covariates of no interest. A significant difference in FA, F1 and RD between no-ECMO and ECMO group is seen in major white matter tracts and subcortical white matter in gyri leading to the conclusion that these differences are driven by alterations in axon coherence. Additionally, we examine individual diffusion measures by looking at masks from 50 brain regions taken from a paediatric brain atlas. We find left anterior corona radiata, left and right corpus callosum (genu, body and splenium), left and right crus of fornix, left anterior limb of internal capsule, left anterior commissure, left tapetum and right uncinate fasciculus to have significantly different means in no-ECMO compared to ECMO group which matches the reports of neuropsychological delays found in behavioural tests. To conclude, analysing diffusion measures at an early stage of life serves as a good tool to detect structural white matter changes in survivors of neonatal ECMO treatment like lacking axon coherence in fibre bundles which develop early in life. The advantage of DWI lies in looking only at the neurobiology, e.g. white matter integrity. Compared to neuropsychological testing, DWI in this age range is a very time-efficient method which does not depend on the child’s active participation. Additional targeted training could help to mitigate the neurodevelopmental deficits ECMO survivors face later in life.

## Introduction

Extracorporeal membrane oxygenation (ECMO) is a life-saving technology for critically ill patients suffering from severe respiratory and/or cardiac failure, applied when maximal conventional therapy has failed. In 1975, neonatal ECMO was – for the first time – successfully administered by Bartlett and colleagues while treating a 1-day-old neonate after failing conventional therapies [1]. According to the 2022 Extracorporeal Life Support Organization (ELSO) registry report [2], ECMO treatments are distributed across adult (54.5 %), pediatric (18.9 %) and neonatal (26.6 %) patient groups – resulting in over 45,000 neonates with ECMO applied worldwide from 1989 to 2022. The majority of these neonates (73.0 %) suffered from pulmonary problems, mostly from congenital diaphragmatic hernia (CDH) which occurs in approximately 1 out of 3,500 - 4,000 live births [3]. A defect of the diaphragm leads to herniation of abdominal organs into the thoracic cavity impacting the growth and development of the lungs so that they remain small and underdeveloped. With the implementation of ECMO treatment, the survival rates after CDH repair – even in neonates with severe CDH – were improved [4]. Despite technological enhancements and increasing clinical practice, significant morbidities among ECMO survivors with or without CDH – besides pulmonary problems – are neurologic complications, with the former showing even more neurodevelopmental problems during childhood [5; 6; 7]. In this study, we combined CDH and ECMO because it is not possible to investigate the effect of ECMO treatment in neonates only, i.e., without underlying disease.

In the neonatal age group, neuronal plasticity is still very dynamic: cortex and basal ganglia develop rapidly, central motor pathways are shaped establishing new cortico-thalamic connections, as well as eliminating old ones [8]. A severe respiratory failure at this time in life interrupts these processes and affects developmental plasticity by modifying not only neurotransmission, but also cellular signalling and neural connectivity/function, leading to regrowth of axons innervating the wrong targets. Although it was shown that neonatal ECMO survivors have an overall average and stable IQ from 2 to 5 to 8 years of age [9; 10], in many follow-up programs, intelligence remains the primary outcome measure in these children [10]. But only 5 % to 10 % of neonatal ECMO survivors suffer from severe neurologic complications, about 90 % of ECMO survivors are at risk for subtler long-term neurodevelopmental problems [9; 11]. Among the neurodevelopmental delays reported for CDH survivors with or without ECMO treatment are verbal, visuo-spatial and working memory problems [11], motor dysfunction as well as sensorineural hearing loss [12; 13]. But it was also shown that no-ECMO treated CDH children have a better prognosis and are less likely to experience co-morbidities that impact on long-term outcome than their ECMO counterparts [13; 14]. On the other hand, a lot of studies which looked at clinical parameters and their correlation to neurodevelopmental outcome later in life did not find any significant associations: neither in CDH survivors, nor in CDH survivors treated with ECMO (see for example 11; 15; 16; 17).

Based on these results, we decided to look only at differences in brain structure. To determine neurodevelopmental delay in ECMO survivors, usually only neuropsychological assessment is used. However, it would be beneficial to identify children who are at higher risk for neurodevelopmental delay later in life as early as possible relying on information extracted directly from the brain. Van den Bosch et al. [18] found cortical thickness and global brain volumes in 8- to 15-year-old neonatal ECMO survivors, despite verbal memory problems, to be similar to healthy controls. They suppose that ECMO survivors suffer from very specific or subtle brain injuries that may not be identifiable using only high resolution structural magnetic resonance imaging (MRI). Therefore, we used diffusion weighted imaging (DWI) to detect alterations in microstructural characteristics in brain white matter (WM) such as WM integrity. WM integrity is undergoing rapid development in the neonatal period due to the dynamic plasticity, so CDH survivors with or without ECMO treatment may show different WM alterations. Moreover, WM integrity has been associated with neuropsychological outcome [19] and behaviour [20].

This study aims to assess whether ECMO treatment associates with WM alterations in 2-year-old CDH survivors and whether these are relatable to the long-term neuropsychological deficits observed in these children later in life. Up-to-now, there are only four studies investigating ECMO survivors by means of DWI: one compared normal age-matched neonates with survivors of hypoxic–ischemic encephalopathy and ECMO [21], one looked at CDH survivors with or without ECMO at school-age [22] and the other two looked at ECMO survivors at school-age [23; 24] – all with limited brain regions analysed. We decided to focus on 2-year-olds when lung growth is already in the middle of the alveolar stage and the recovery process from ECMO treatment no longer influences DWI parameter measurement. On the other hand, at 2 years of age, the children are not yet too old to benefit from additional therapies or exercises. Therefore, examining WM integrity in this age cohort may reveal more clearly side effects of ECMO treatment in the long run. We compare whole-brain WM integrity extracted from DWI in CDH survivors after ECMO treatment to a no-ECMO CDH control group. Then, we analyse 50 distinct brain regions including major white matter tracts and subcortical white matter in gyri. We expect to find WM alterations in ECMO survivors, specifically in areas associated with working memory, attention and motor function.

## Materials & Methods

### Patients

This study incorporates 56 children (mean age at measurement: 25.88 months, SD = 5.52 months) suffering from CDH, which were investigated in our institution according to the local follow-up program including MRI [25; 26]. All children born between 2011 and 2017 in whom the conventional MRI and DWI measurements were of diagnostic quality were considered for inclusion. The children were delivered at our institution with the antenatal diagnosis of CDH. Because of respiratory failure, 18 (7 girls, 11 boys) out of the 56 children received ECMO treatment (mean duration of ECMO: 9.41 days, SD = 3.18 days). ECMO treatment was initiated according to the recommendations suggested by the CDH EURO Consortium Consensus and ELSO [27]. Within the ECMO group, nine out of 18 children (4 girls, 5 boys) received surgical reconstruction of the right common carotid artery (rCCA), the other nine children (3 girls, 6 boys) received ligation. Thirty-eight children (14 girls, 24 boys) served as “control group” suffering from CDH, but without ECMO treatment applied. As extensively described elsewhere, all children receiving ECMO were treated according to our postnatal management schedule [28, 29]. In the MRI sessions, an additional diffusion measurement – designed especially for the study purpose – was included. The children who received ECMO treatment stayed longer in hospital (mean duration: 75.18 days; SD = 38.00 days) than those without ECMO treatment (mean duration: 33.39 days, SD = 23.23 days). Written informed consent was obtained by the children’s parents. The study was approved by the local ethics committee (Ethikkommission II der Universität Heidelberg, Medizinische Fakultät Mannheim), all research was performed in accordance with relevant guidelines/regulations and with the Declaration of Helsinki.

### DWI data acquisition

During DWI measurement, all children were sedated by intravenous administration of Propofol and continuously monitored by an anaesthesiologist. The measurement was performed on a 3 T whole-body MRI scanner (Magnetom TimTrio, Siemens Healthineers, Erlangen, Germany) using a 12-channel head coil. For acquisition of DWI, a spin-echo echo planar imaging sequence (TR = 8400 ms, TE = 84 ms, FoV = 192 × 192 mm^2^, BW = 1930 Hz/px) was used. Forty-seven slices (voxel size = 2.0 mm^3^, no gap) were acquired in interleaved slice order using GRAPPA acceleration factor 2 and two averages. Diffusion weighting was performed in multi-directional diffusion weighting mode along 30 non-collinear directions with b = 1000 s/mm^2^. Additionally, a single non-diffusion weighted volume (b = 0 s/mm^2^) was acquired with every average. Acquisition time was 5:58 minutes.

### DWI data processing and statistics

For DWI analyses, data were denoised first using MRtrix3 [30] (*dwidenoise*) and then pre-processed using FMRIB software library v 6.0.2 [31] running on Ubuntu 18.04.3 LTS. After correction of eddy current distortions (*eddy*) and subject movement, we used *bet* routine with centre-of-gravity (-c) option to extract brain masks and adjusted fractional intensity threshold (f = 0.2) and vertical gradient in fractional intensity threshold (g = −0.1) option, accordingly [32]. To calculate fractional anisotropy (FA) from eigenvector maps, *dtifit* routine with weighted least-squares regression was used. From these results, radial diffusivity (RD) values were determined. A modified version of Tract-Based Spatial Statistics (TBSS) [33] was used for voxelwise statistical analyses. All scripts were changed as to register all children’s FA data onto the Johns Hopkins University MRI/DTI Pediatric Brain Atlas for 2-year-olds (https://cmrm.med.jhmi.edu/) [34] in a first step, afterwards a mean FA image was created and thinned to a mean FA skeleton which represents the centres of all tracts common to the group. The projection of every child’s aligned FA data onto the mean FA skeleton was fed into voxelwise cross-subject statistics [35] with 5000 permutations of the data using age and sex as covariates of no interest. For analyses of RD data, we used a TBSS script (*tbss_non_FA*) adapted for 2-year-olds by changing all references to standard space template images and masks. Furthermore, crossing-fibre parameters were estimated using *bedpostX* routine with Rician noise option and 3 fibres modelled per voxel. The resulting fibre orientation and partial volume fraction estimates were analysed using a TBSS script (*tbss_x*) [36] for crossing fibres adapted for 2-year-olds by changing all references to standard space template images and masks. All resulting projections were, again, fed into whole-brain voxelwise cross-subject statistics using age and sex as covariates of no interest. All TBSS analysis results were acquired using Threshold-Free Cluster Enhancement [37] and are fully corrected for multiple comparisons using family-wise error (FWE) rate.

For statistical correlation analyses, mean values of diffusion measures were extracted from the corresponding parameter maps using masks from the Johns Hopkins University Pediatric Brain Atlas parcellation file. The masks were chosen according to Johns Hopkins University ICBM-DTI-81 WM atlas [38] for adults included in FSL (for more details about masks selected see Table 1). We did this to increase comparability to later studies when our children were older, or to studies that include only older children/adolescents. Although much of brain development occurs during the first few years of life, this process continues well beyond infancy [57] and throughout adolescence.

**Table 1:** Labels of masks used in analyses and corresponding names of brain regions. The masks were selected from the Lookup Table of the Johns Hopkins University MRI/DTI Pediatric Brain Atlas for 2-year-olds corresponding to masks used in ICBM-DTI-81 WM atlas. R/L denotes right/left hemisphere, respectively.

| Labels |  | Region |
| --- | --- | --- |
| *ICBM-DTI-81* | *JHU_pediatric24_SS* |  |
| ant_CC | Genu_CC_R | Genu of corpus callosum R/L |
|  | Genu_CC_L |  |
| CC | Body_CC_R | Body of corpus callosum R/L |
|  | Body_CC_L |  |
| post_CC | Splenium_CC_R | Splenium of corpus callosum R/L |
|  | Splenium_CC_L |  |
| R_cer_peduncle | CP_R | Cerebral peduncle R |
| L_cer_peduncle | CP_L | Cerebral peduncle L |
| R_alic | ALIC_R | Anterior limb of internal capsule R |
| L_alic | ALIC_L | Anterior limb of internal capsule L |
| R_plic | PLIC_R | Posterior limb of internal capsule R |
| L_plic | PLIC_L | Posterior limb of internal capsule L |
| R_Ant_cor_rad | ACR_R | Anterior corona radiata R |
| L_Ant_cor_rad | ACR_L | Anterior corona radiata L |
| R_Sup_cor_rad | SCR_R | Superior corona radiata R |
| L_Sup_cor_rad | SCR_L | Superior corona radiata L |
| R_Post_cor_rad | PCR_R | Posterior corona radiata R |
| L_Post_cor_rad | PCR_L | Posterior corona radiata L |
| R_ilf | SS_R | Sagittal stratum (include inferior longitudinal fasciculus and inferior fronto-occipital fasciculus) R |
| L_ilf | SS_L | Sagittal stratum (include inferior longitudinal fasciculus and inferior fronto-occipital fasciculus) L |
| R_unc | EC_R | External capsule R |
| L_unc | EC_L | External capsule L |
| R_cing | CGC_R | Cingulum (cingulate gyrus) R |
| L_cing | CGC_L | Cingulum (cingulate gyrus) L |
| R_cing_hipp | CGH_R | Cingulum (hippocampus) R |
| R_cing_hipp | CGH_L | Cingulum (hippocampus) L |
| R_hipp | Fx/ST_R | Fornix (cres) / Stria terminalis (cannot be resolved with current resolution) R |
| L_hipp | Fx/ST_L | Fornix (cres) / Stria terminalis (cannot be resolved with current resolution) L |
| R_slf | SLF_R | Superior longitudinal fasciculus R |
| L_slf | SLF_L | Superior longitudinal fasciculus L |
| R_unc_needsTBC | UNC_R | Uncinate fasciculus R |
| L_unc_needsTBC | UNC_L | Uncinate fasciculus L |
| R_tapetum | Tapetum_R | Tapetum R |
| L_tapetum | Tapetum_L | Tapetum L |
|  | AnteriorComm_L | Anterior commissure L |
|  | AnteriorComm_R | Anterior commissure R |
|  | AnsaLenticularis_L | Ansa lenticularis L |
|  | AnsaLenticularis_R | Ansa lenticularis R |
|  | OpticTract_L | Optic tract L |
|  | OpticTract_R | Optic tract R |
|  | LenticularFasc_L | Lenticular fasciculus L |
|  | LenticularFasc_R | Lenticular fasciculus R |
|  | SubstriatumWM_L | Sub-striatum white matter L |
|  | SubstriatumWM_R | Sub-striatum white matter R |
|  | ML_L | Medial lemniscus L |
|  | ML_R | Medial lemniscus R |
|  | SFO_L | Superior fronto-occipital fasciculus L |
|  | SFO_R | Superior fronto-occipital fasciculus R |
|  | IFO_L | Inferior fronto-occipital fasciculus L |
|  | IFO_R | Inferior fronto-occipital fasciculus R |
|  | PTR_L | Posterior thalamic radiation (include optic radiation) L |
|  | PTR_R | Posterior thalamic radiation (include optic radiation) R |

We refrained from performing tractography considering the still persisting problems with the tracking algorithms – even in phantoms – as described by Schilling et al. [39]. Furthermore, the atlas-based approach we used here offers a better predictive accuracy than approaches including streamline tractography [40].

For testing of categorical and continuous variables, we performed a *χ^2^* test with α = 0.05 and Welch’s *t*-tests with α = 0.05, respectively, as implemented in MATLAB R2013a (The MathWorks Inc., Natick, MA, USA). For further analyses of FA and RD values taken from each individual mask, we tested for normality in each sample, using the Shapiro-Wilk test [41] with α = 0.05 and concluding that normality could not be assumed. Using the Levene test [42] with α = 0.05, we tested whether variances could be assumed equal for both groups. We concluded that this was not the case. Therefore, the nonparametric Brunner-Munzel test [43] with α = 0.025 corrected for multiple inference (Bonferroni correction) was chosen for further analysis. This test is specifically designed to compare the location of two samples in the possible presence of unequal variances. It does not assume normality [44].

## Results

Due to excessive head motion (> 2 mm in translation and 2° in rotation) or application of wrong measurement protocols, seven children - one from ECMO group and six from no-ECMO group - were excluded. The sample size for further analyses is now 49.

All conventional MRI measurements were checked for clinical evidence by an experienced paediatric radiologist. Because we have limited clinical information of the children due to lack of a digital patient management system, additional data – including data on neurodevelopmental assessment – is not available. In the two groups, the clinical parameters relevant for decision of ECMO treatment (see guidelines [27] for details) are significantly different because otherwise the children in the ECMO group would not have needed ECMO.

We performed Welch’s *t*-tests on age and duration of stay in hospital. There is no significant difference between ECMO group and no-ECMO group in terms of age (t(28.262) = 1.523, p = 0.139). There is a significant difference between ECMO group and no-ECMO group in terms of duration of stay in hospital (t(22.526) = 4.146, p < 0.001) which is caused by the need for ECMO treatment. Furthermore, we performed a *χ^2^* test on sex. There is no significant difference between ECMO group and no-ECMO group in terms of sex (*χ^2^*(1, *N* = 49) = 0.030, p = 0.862).

Within the ECMO group, there is no significant difference in diffusion measures between children receiving ligation and those receiving surgical reconstruction of rCCA. For further analyses, we merged the two ECMO subgroups.

In the whole-brain voxelwise analysis, we find significantly higher (p_FWE_ < 0.05) FA values in no-ECMO group compared to ECMO group (Fig. 1, upper panel) in centres of the following WM tracts: left and right anterior corona radiata, left and right corpus callosum (genu, body and splenium), left and right posterior thalamic radiation, left superior corona radiata, left and right middle occipital gyrus, right crus of fornix, left precuneus, left superior parietal gyrus and left superior occipital gyrus, as well as right inferior frontal gyurs and right middle frontal gyrus.

**Figure 1:**
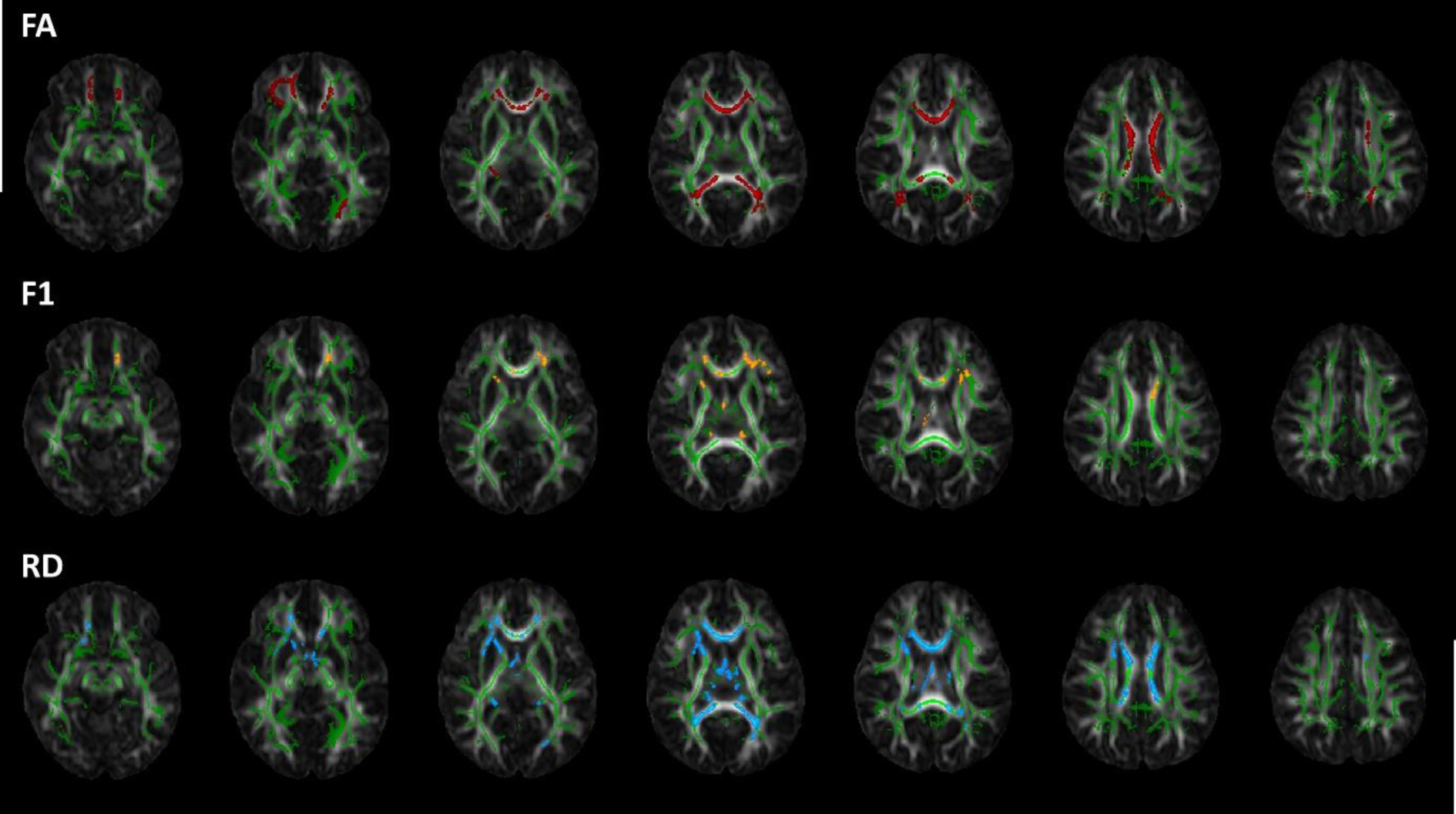
Voxelwise statistical analyses results of fractional anisotropy (FA), first fibre partial volume fraction estimate (F1) and radial diffusion (RD). The mean FA skeleton (green) is overlaid on the FA image. Statistically significant differences in the contrast no-ECMO > ECMO are shown for FA (red) and F1 (yellow). Statistically significant differences in the contrast ECMO > no-ECMO are shown for RD (blue). The slices range from z = 50 to z = 94 in standard space and are displayed in neurological orientation.

Because FA changes in crossing fibre regions are difficult to interpret, we additionally performed a crossing fibre analysis setting three fibres per voxel. In the whole-brain voxelwise analysis, we find significantly higher (p_FWE_ < 0.05) first fibre partial volume fraction estimates (F1 - Fig. 1, middle panel) in no-ECMO group compared to ECMO group in the centres of the following WM tracts: left and right anterior corona radiata, left and right corpus callosum (genu, body and splenium), left and right superior corona radiata, right crus of fornix, left posterior thalamic radiation, right anterior limb of internal capsule, left external capsule, left cingulum (cingulate gyrus), left and right thalamus, left and right midbrain, left inferior frontal gyrus, as well as right superior frontal gyrus.

There are eleven WM tracts with higher FA and higher F1 values in no-ECMO group compared to ECMO group. In these brain regions, higher F1 values indicate higher orientation of fibres and fibre coherence. To support this finding, we further analysed RD values which give insight into radial diffusion. A whole-brain voxelwise analysis revealed significantly higher (p_FWE_ < 0.05) RD values in ECMO group compared to no-ECMO group in the centres of the following WM tracts: right anterior corona radiata, left and right corpus callosum (genu, body and splenium), left and right anterior commissure, left posterior thalamic radiation, left crus of fornix, left and right thalamus and left superior corona radiata (Fig. 1, lower panel).

In total, there are ten WM tracts with higher FA, higher F1 and lower RD values in no-ECMO group compared to ECMO group: right anterior corona radiata, left and right corpus callosum (genu, body and splenium), right crus of fornix, left posterior thalamic radiation and left superior corona radiata.

Because whole-brain voxelwise statistical analyses compare the group means by projecting diffusion measures onto the centres of all WM tracts common to the group, we additionally performed atlas-based analyses by looking at the individual FA and RD values per mask. In Fig. 2, the mean FA values per mask for both groups are shown. It is obvious that all mean FA values of the no-ECMO group are higher than the corresponding mean FA values of the ECMO group. To statistically examine the distribution of mean FA values of ECMO and no-ECMO group, we performed Brunner-Munzel tests.

**Figure 2:**
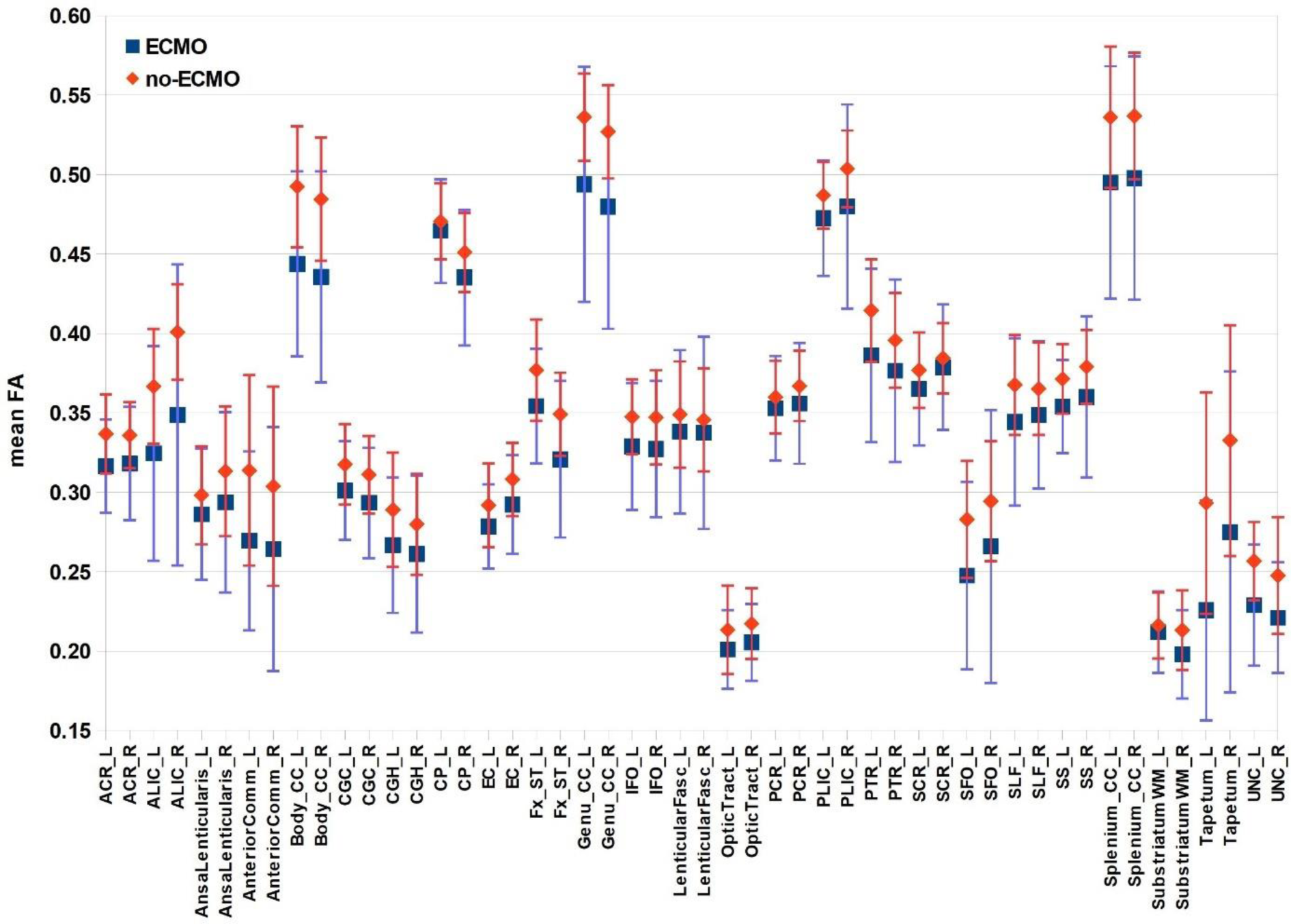
Mean fractional anisotropy (FA) values of white matter atlas masks. The mean FA values per mask and their standard deviations are plotted for both groups (ECMO, no-ECMO). The masks were taken from Johns Hopkins University MRI/DTI Pediatric Brain Atlas for 2-year-olds (see Table 1).

We find a significant difference in no-ECMO > ECMO in mean FA values in the following masks: left anterior corona radiata (W^BF^(33.355) = 2.717, p = 0.005), left and right anterior limb of internal capsule (left: W^BF^(34.366) = 3.744, p < 0.001; right: W^BF^(25.533) = 2.849, p = 0.004), left anterior commissure (W^BF^(35.507) = 2.291, p = 0.014), left and right genu of corpus callosum (left: W^BF^(25.046) = 3.183, p = 0.002; right: W^BF^(28.129) = 3.939, p < 0.001), left and right body of corpus callosum (left: W^BF^(29.803) = 4.210, p < 0.001; right: W^BF^(22.566) = 3.297, p = 0.002), left and right splenium of corpus callosum (left: W^BF^(42.466) = 3.772, p < 0.001; right: W^BF^(35.855) = 3.608, p < 0.001), right external capsule (W^BF^(38.863) = 2.047, p = 0.024), left and right crus of fornix (left: W^BF^(37.057) = 2.794, p = 0.005; right: W^BF^(23.449) = 2.467, p = 0.011), left tapetum (W^BF^(34.655) = 3.801, p < 0.001) and left and right uncinate fasciculus (left: W^BF^(40.772) = 3.014, p = 0.002; right: W^BF^(40.340) = 2.995, p = 0.002).

In Fig. 3, the mean RD values per mask for both groups are shown. Almost all mean RD values of the ECMO group are higher than the corresponding mean RD values of the no-ECMO group. Again, to statistically examine the distribution of mean RD values of ECMO and no-ECMO group, we performed Brunner-Munzel test. In mean RD values, we find a significant difference in ECMO > no-ECMO in the following masks: left anterior corona radiata (W^BF^(30.382) = −2.778, p = 0.005), left anterior limb of internal capsule (W^BF^(30.246) = −2.565, p = 0.008), right ansa lenticularis (W^BF^(32.436) = −2.696, p = 0.006), left and right anterior commissure (left: W^BF^(32.426) = −3.204, p = 0.002; right: WBF(29.027) = −3.306, p = 0.001), left and right cerebral peduncle (left: WBF(31.959) = −2.482, p = 0.009; right: W^BF^(26.033) = −3.323, p = 0.001), left and right crus of fornix (left: W^BF^(32.333) = −3.556, p = 0.001; right: W^BF^(26.643) = −2.881, p = 0.004), left and right genu of corpus callosum (left: W^BF^(34.521) = −4.563, p < 0.001; right: W^BF^(31.269) = −4.253, p < 0.001), left and right body of corpus callosum (left: W^BF^(37.409) = −4.701, p < 0.001; right: W^BF^(34.064) = −4.208, p < 0.001), left and right splenium of corpus callosum (left: W^BF^(46.098) = −5.691, p < 0.001; right: W^BF^(40.895) = −4.198, p < 0.001), left and right superior fronto-occipital fasciculus (left: W^BF^(43.625) = −5.262, p < 0.001; right: W^BF^(25.313) = −2.971, p = 0.003), left and right posterior corona radiata (left: W^BF^(37.195) = −2.429, p = 0.010; right: W^BF^(29.115) = −3.220, p = 0.002), left optic tract (W^BF^(43.916) = −2.163, p = 0.018), left posterior thalamic radiation (W^BF^(28.883) = −2.850, p = 0.004), left tapetum (W^BF^(41.641) = −4.100, p < 0.001) and right uncinate fasciculus (W^BF^(42.510) = −2.601, p = 0.006).

**Figure 3:**
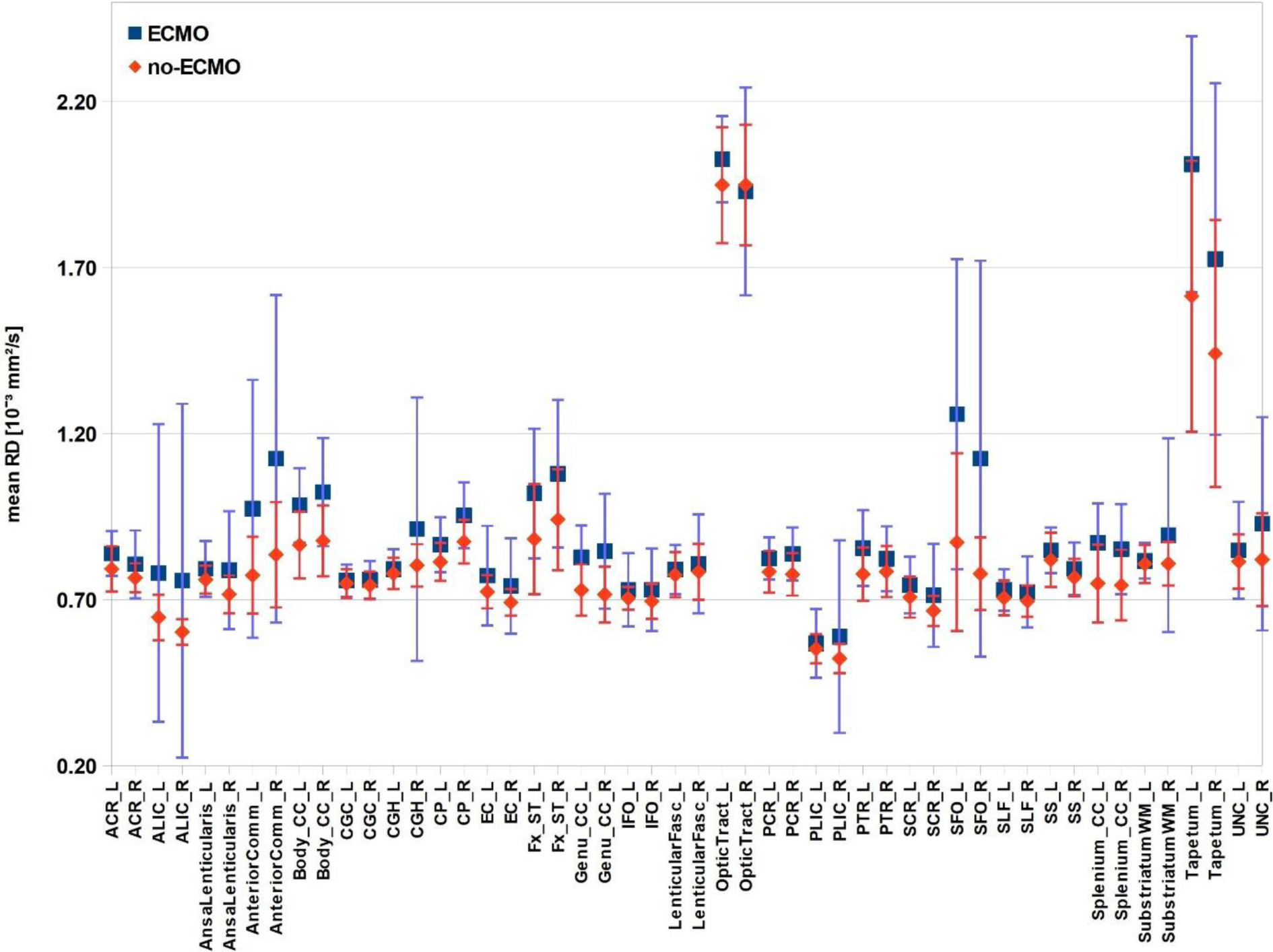
Mean radial diffusivity (RD) values of white matter atlas masks. The mean RD values of the masks and their standard deviations are plotted for both groups (ECMO, no-ECMO). The masks were taken from Johns Hopkins University MRI/DTI Pediatric Brain Atlas for 2-year-olds (see Table 1).

In total, there are 13 brain masks with significantly higher FA and significantly lower RD values in no-ECMO group compared to ECMO group: left anterior corona radiata, left anterior limb of internal capsule, left anterior commissure, left and right corpus callosum (genu, body and splenium), left and right crus of fornix, left tapetum and right uncinate fasciculus (see Fig. 4. for a visual presentation).

**Figure 4:**
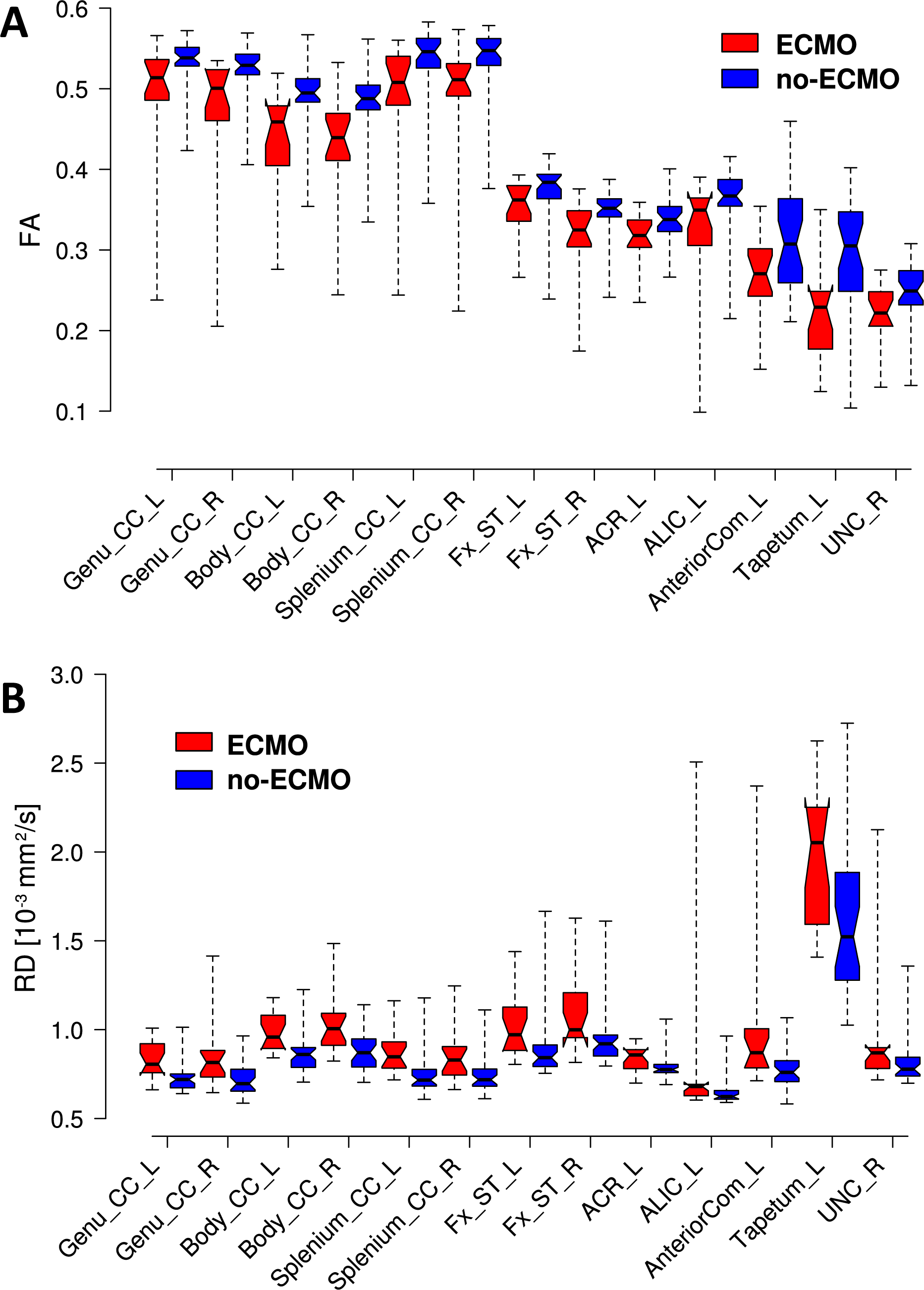
Significantly different fractional anisotropy (FA) and radial diffusivity (RD) values of white matter atlas masks. **A:** The FA values of the significantly different masks are plotted for both groups (ECMO, no-ECMO). **B:** The RD values of the significantly different masks are plotted for both groups (ECMO, no-ECMO). Centre lines show the group medians; notches correspond to 95% confidence interval of median; box limits indicate the 25^th^ and 75^th^ percentiles; whiskers extend to minimum and maximum values. The masks were taken from Johns Hopkins University MRI/DTI Pediatric Brain Atlas for 2-year-olds (see Table 1).

The boxplots visualise very well the median and spread of values within each group; in all 13 brain masks, the medians of FA values in no-ECMO group are higher compared to ECMO group (Fig. 4 A) and the spread of FA values is smaller. On the other hand, the medians of RD in the same regions are lower (Fig. 4 B) with the spread of RD values again remaining smaller in no-ECMO group compared to ECMO group. The numerical data of all medians and interquartile ranges are listed in Table 2.

**Table 2:** Median (Mdn) and interquartile range (IQR) values of fractional anisotropy (FA) and radial diffusivity (RD) from brain regions shown in Fig. 4 for both groups: ECMO and no-ECMO. The masks were selected from the Lookup Table of the Johns Hopkins University MRI/DTI Pediatric Brain Atlas for 2-year-olds.

| Name of Atlas<br>Mask | Mdn FA (IQR) | | Mdn RD (IQR) [$10^{-3}$ mm<sup>2</sup>/s] | |
| --- | --- | --- | --- | --- |
|  | ECMO | no-ECMO | ECMO | no-ECMO |
| Genu_CC_L | 0.51 (0.49 – 0.54) | 0.54 (0.53 – 0.55) | 0.81 (0.76 – 0.92) | 0.72 (0.67 – 0.75) |
| Genu_CC_R | 0.50 (0.46 – 0.52) | 0.53 (0.52 – 0.54) | 0.81 (0.73 – 0.88) | 0.70 (0.65 – 0.78) |
| Body_CC_L | 0.46 (0.40 – 0.48) | 0.49 (0.48 – 0.51) | 0.96 (0.89 – 1.08) | 0.86 (0.79 – 0.90) |
| Body_CC_R | 0.44 (0.41 – 0.47) | 0.49 (0.47 – 0.50) | 1.01 (0.91 – 1.09) | 0.87 (0.79 – 0.95) |
| Splenium_CC_L | 0.51 (0.48 – 0.54) | 0.55 (0.53 – 0.56) | 0.85 (0.78 – 0.93) | 0.72 (0.68 – 0.78) |
| Splenium_CC_R | 0.51 (0.49 – 0.53) | 0.55 (0.53 – 0.56) | 0.83 (0.74 – 0.91) | 0.72 (0.68 – 0.78) |
| Fx_ST_L | 0.36 (0.34 – 0.38) | 0.38 (0.36 – 0.39) | 0.97 (0.88 – 1.13) | 0.84 (0.79 – 0.91) |
| Fx_ST_R | 0.32 (0.30 – 0.35) | 0.35 (0.34 – 0.36) | 1.00 (0.96 – 1.21) | 0.92 (0.85 – 0.97) |
| ACR_L | 0.32 (0.30 – 0.34) | 0.34 (0.32 – 0.35) | 0.86 (0.78 – 0.89) | 0.78 (0.76 – 0.80) |
| ALIC_L | 0.35 (0.31 – 0.36) | 0.37 (0.35 – 0.39) | 0.68 (0.63 – 0.69) | 0.62 (0.61 – 0.66) |
| AnteriorCom_L | 0.27 (0.24 – 0.30) | 0.31 (0.26 – 0.36) | 0.87 (0.79 – 1.01) | 0.76 (0.71 – 0.83) |
| Tapetum_L | 0.23 (0.18 – 0.25) | 0.31 (0.25 – 0.35) | 2.05 (1.59 – 2.25) | 1.52 (1.28 – 1.89) |
| UNC_R | 0.22 (0.21 – 0.25) | 0.25 (0.23 – 0.27) | 0.87 (0.78 – 0.90) | 0.78 (0.74 – 0.85) |

## Discussion

We could show that ECMO treatment associates with WM integrity in 2-year-old CDH survivors. DWI measures like FA include effects from many underlying parameters and are thus non-specific. There are many physiological processes that may cause changes in imaging parameters to occur during brain development. Possible microstructural contributions include myelination or changes in axonal density or axon coherence. Concurrent changes in tract volume and water content also influence various imaging parameters. In the whole-brain voxelwise analyses and in almost all WM masks, we clearly see higher FA values (see Fig. 1, upper panel and Fig. 2) and lower RD values (see Fig. 1, lower panel and Fig. 3) in no-ECMO group compared to ECMO group. This may be due to various underlying parameters including axonal density, fibre orientation dispersion, and degree of myelination. Roberts et al. [45] and Friedrich et al. [46] report no correlation between myelin content and FA. But there was a correlation between axon/fibre density and FA. This is also supported by Winston et al. [47] who report a decrease in FA in focal cortical dysplasia in epilepsy patients due to an increase in extracellular space. Furthermore, Stikov et al. [48] found that FA is most sensible to the amount of extra-axonal space which is related to axon density as can be seen in areas with high directional coherence, such as the corpus callosum where FA increases. On the basis of these findings, increases in FA here seem to be merely based on either increase in axon density or axon coherence – when more axons are aligned along the same axis.

Our results show significantly lower FA values in ECMO group in WM tracts associated with high axon coherence, such as the corpus callosum and the fornix. Besides, the first fibre partial volume fraction estimate F1 was lower in ECMO group in these tracts supporting this conclusion. Simultaneously, RD values in the same tracts are significantly higher indicating either less axon density or less axons aligned along the same axis. This suggests that, due to the very dynamic neuronal plasticity at the neonatal stage, children from ECMO group experience neural connectivity which leads to erroneous growth of axons that innervate the wrong targets. For a developing brain, exposure to hypoxic ischemic injury around the time of birth can be considered as a point of vulnerability in neurobiological development, the neurodevelopmental impairments ECMO survivors exhibit later in life match these findings. The most relevant brain regions (significant higher FA and significant lower RD in no-ECMO group – Fig. 4) we found in the individual FA and RD values support the findings on impairment in neurodevelopmental outcome later in life reported by others [11; 12; 15; 16; 17; 23; 24; 49; 50; 51; 52] which are a) motor dysfunction (corpus callosum and anterior limb of internal capsule), b) visuo-spatial problems (corpus callosum and anterior commissure) c) memory problems (crus of fornix and uncinate fasciculus) and d) attention problems (anterior corona radiata and anterior commissure). The three parts of the corpus callosum (genu, body and splenium) connect the two cerebral hemispheres and are known to be involved in movement control, cognitive functions and vision. The anterior limb of the internal capsule relays motor and sensory information with ascending and descending fibres between the cerebral cortex and the pyramids of the medulla [53]. The crus of fornix acts as the major output tract of the hippocampus connecting it to various subcortical structures like the mammillary bodies and the anterior nucleus of thalamus. It is also a critical component of the Papez circuit which is involved in learning and memory, emotion and social behaviour. The uncinate fasciculus connects parts of the limbic system and plays a role in memory encoding/memory retrieval and regulation of emotion. The anterior corona radiata is part of the limbic-thalamo-cortical circuitry and consists of afferent and efferent fibres that connect the cerebral cortex and the brain stem. Both are involved in sensation and motor function, and the corona radiata connects motor and sensory nerve pathways between these structures. Niogi et al. [54] provide evidence that microstructural integrity of the anterior corona radiata modulates executive attention. The anterior commissure plays a significant role in visual processing and memory, among others, as was shown by selectively inactivating the anterior commissure in monkeys (see review by Fenlon *et al*. [55]).

There was no significant difference in diffusion measures between ligation and rCCA within the ECMO group, which is in line with Duggan et al. [56] who were able to show that there appear to be no significant differences in the incidence of brain lesions in patients who undergo carotid repair at time of decannulation from ECMO compared to those undergoing ligation. Likewise, clinical parameters such as persistent pulmonary hypertension, arterial blood gas values, pre- and postductal oxygen saturation, cardiovascular dysfunction and many others are not correlated with adverse neurodevelopmental delay [15]. Gestational age is not an important factor of developmental outcomes after ECMO as well [16]. The study by Madderom et al. [11] assessed the influence of medical variables on the neuropsychologic domains on which adolescent survivors of neonatal ECMO treatment showed impaired performance and did not find any of these variables to be a significant predictor. A fourth study identified ventilator time/need for ECMO as the only independent predictor of motor problems at age 1 [17]. Danzer et al. [15] investigated the influence of need and timing of ECMO in relation to CDH repair on short-term neurodevelopmental outcomes in children of 2 years of age. They did not find any association of duration of ECMO with a higher likelihood of adverse cognitive, language or motor outcome or increased risk of neuromuscular hypotonicity. Furthermore, if children in need of neonatal ECMO were repaired early or late in the ECMO course did not have an impact on their neurodevelopmental outcome.

The advantage of DWI lies in its freedom from bias while looking only at the neurobiology, e.g. WM integrity. DWI is a very efficient and reliable method which does not depend on the child’s active participation. Compared to other DWI studies where only limited brain regions (six regions [22; 23; 24] or nine regions [21]) were selected, we performed whole-brain TBSS analyses based on a paediatric atlas for 2-year-olds. Ten WM tracts show a significant difference in all three diffusion measures (FA, F1, RD). We then selected the corresponding atlas masks plus those equivalent to the masks given in Johns Hopkins University ICBM-DTI-81 WM atlas for adults shipped with FSL for further analyses on individual level (see Table 1) and find significantly different group means in thirteen masks that match the results of the neurodevelopmental assessments reported before (see above).

To conclude, analysing diffusion measures such as FA, F1 and RD serves as a tool to detect early structural WM changes in survivors of neonatal ECMO treatment. The differences lie in lacking axon coherence in fibre bundles which develop early in life. As these children are at risk for developing cognitive disorders during childhood which persist into adolescence [11; 12], we recommend focused screening to support therapeutic strategies. Schiller et al. [24] have already shown that training-induced changes in WM microstructure, e.g. after Cogmed Working-Memory Training, are possible and associated with better verbal working-memory. This procedure should also be applicable to the other areas of neurodevelopmental impairment seen later in life because, although much of brain development occurs during the first few years of life, this process continues well beyond infancy [57] and throughout adolescence. Targeted training can therefore help to mitigate the neurodevelopmental deficits ECMO survivors face later in life.

## Data Availability

All data produced in the present study are available upon reasonable request to the authors.

## Author Contributions

MR performed the data and statistical analyses and wrote the paper. JF pre-sorted and sifted the data and participated to the statistical analysis. TS participated to the coordination of the study, recruited children and provided the clinical expertise. MW supervised MR acquisition, provided radiological expertise and checked for clinical evidence. FZ helped in the interpretation of the results and participated to the paper writing.

## Conflict of Interest

The authors have indicated they have no potential conflicts of interest to disclose.

## Abbreviations

ECMO: extracorporeal membrane oxygenation
DWI: diffusion weighted imaging
WM: white matter
FA: fractional anisotropy
RD: radial diffusivity
TBSS: Tract-Based Spatial Statistics
TFCE: Threshold-Free Cluster Enhancement

## Notes

### Competing Interest Statement

The authors have declared no competing interest.

### Funding Statement

This work was supported by Deutsche Forschungsgemeinschaft (grant number: DFG 397806429).

### Author Declarations

Ethikkommission II der Universitaet Heidelberg, Medizinische Fakultaet Mannheim, gave ethical approval for this work.

